# Network meta-analysis of oral Chinese patent medicine combined with SSRI for the treatment of depressive disorder in special population

**DOI:** 10.1101/2025.09.04.25335139

**Authors:** Fuqing Ren, Xueyan Liu, Jinbang Wang, Lixian Cui, Jiangyan Wei, Bin Li

**Author notes:** Corresponding author: Bin Li. No.11 Third Ring East Road, Chaoyang District, Beijing 100029, China (Xueyan Liu). No. 6 Fangzhuang Fangxingyuan 1st District, Fengtai District, Beijing 100078, China (Lixian Cui). No. 23 Meishuguan Houjie, Dongcheng District, Beijing 100010, China (Bin Li). Fuqing Ren and Xueyan Liu contributed equally to this work. **Competing interests:**The authors have declared that no competing interests exist. **Data Availability Statement:**All data are in the manuscript and/or Supporting information files.

## Abstract

**Objective:** Investigating the therapeutic efficacy and safety of combined oral Chinese patent medicine with selective serotonin reuptake inhibitors (SSRIs) in special Homo sapiens populations with depressive disorders.

**Methods:** China National Knowledge Infrastructure(CNKI), Chinese Biomedical Literature Database(CBM), Chinese Academic Journal Database(CSPD), Chinese Science and Technology Journal Database(CCD), EMbase, Pubmed,Cochrane Library, and Web of Science were retrieved, and ClinicalTrials.gov was searched from the establishment of the database to October 2023. All included Rized controlled trials were assessed for quality using the Cochrane systematic evaluation manual.Network Meta-analysis was performed using the Stata 17.0 software.

**Results:** 36 studies were included, and the total sample size included 3110 cases, including 1556 cases in the test group and 1554 cases in the control group. A total of Chinese patent medicines were included: Shugan Jieyu capsule, Xiaoyaosan , Wuling capsule, and Morinda officinalis oligose. The results of the network Meta-analysis showed that. Compared to antidepressant treatment with SSRI alone, Adolescent depressive disorder: In improving clinical efficiency, Wuling capsule combined with SSRI ranked the highest; In terms of improving the HAMD score, Xiaoyaosan ranked the highest; Geriatric depressive disorder: In improving clinical efficiency, Shugan Jieyu capsule combined with SSRI ranked the highest; In terms of improving the HAMD score, the Shugan Jieyu capsule combined with SSRI ranked the highest; Postpartum depression disorder: In improving clinical efficiency, the Shugan Jieyu capsule combined with SSRI ranked the highest; In terms of improving the HAMD score, the Shugan Jieyu capsule combined with SSRI ranked the highest; Perimenopausal depression disorder: In improving clinical efficiency, the Xiaoyaosan combined with SSRI ranked the highest; In terms of improving the HAMD score, the Wuling capsule combined with SSRI ranked the highest;

**Conclusion:** On the basis of SSRI, the addition of Chinese patent medicine can improve the effectiveness and safety of clinical treatment. It is suggested that more high-quality RCT studies of Chinese patent medicine for depression disorders, especially in adolescent depression disorder, postpartum depression disorder, perimenopausal depression disorder, are conducted to provide stronger evidence.

**Trial registration:** This systematic review protocol has been registered on PROSPERO database(www.crd.york.ac.uk/prospero/) with number (CRD42022364242).

## Introduction

Depressive disorder refers to a group of mood disorders caused by various factors, characterized by prominent and persistent low mood as the main clinical feature. It is one of the most common mental disorders [1] .Epidemiological surveys show that the annual prevalence rate of depressive disorder in China is 3.6%, and the lifetime prevalence rate is 6.8% [2] . It is estimated that by 2030, depressive disorder will surpass tumors and cardiovascular and cerebrovascular diseases to become the leading cause of global disease burden [3] . The occurrence of depressive disorder is also closely related to gender and age, among which children and adolescents, the elderly, and women are special populations prone to depressive disorder.The prevalence rate of depressive emotions among Chinese adolescents is as high as 15.4%, with a total detection rate of depressive emotions reaching 28.4% [4] [5] . Depressive disorder has become the primary cause of disability and morbidity among adolescents aged 10-19 years. Selective Serotonin Reuptake Inhibitors (SSRIs) can be used for depressive disorder in children and adolescents [6] . Geriatric depression is often misunderstood as a physiological aging process, so it has not received systematic diagnosis and treatment for many years. The prevalence rate of geriatric depression in China is 15.9%, among which 36.4% of patients have mild cognitive impairment [7] . In terms of pharmacotherapy, SSRIs are the first-choice drugs [6] . Postpartum depression commonly occurs within 2 weeks after childbirth, and its symptoms can last for up to 1 year. The incidence of postpartum depression among women of childbearing age in China is 14.7%, while the incidence among elderly parturient women can be as high as 36.9% [8] ,[9] . Currently, cognitive behavioral therapy is the most commonly used method in clinical practice for treating mild to moderate postpartum depression, and SSRIs are the first choice for severe postpartum depression [6] . Perimenopausal depressive disorder mostly occurs in women aged 45-55 years. Statistics show that the incidence rate of perimenopausal depressive disorder among Chinese women is 46% [10] ,[11] . For mild cases, psychotherapy can be provided; for moderate to severe cases, combined use of SSRIs is recommended, and hormone replacement therapy can also effectively alleviate depressive symptoms [6] .SSRIs have definite effects in clinical practice and are often recommended as first-line drugs of choice. However, they still have shortcomings such as obvious adverse reactions, withdrawal reactions, and drug resistance, leading to poor patient compliance [12] . Chinese patent medicines have advantages such as convenient administration, portability, and easy storage. Surveys show that approximately 70% of Chinese patent medicines are prescribed by Western medicine physicians Meanwhile, the combination of traditional Chinese medicines and chemical drugs may enhance efficacy and reduce side effects [14] . Nevertheless, due to the wide variety of such combinations and the lack of randomized controlled trials comparing different drugs, it brings inconvenience to clinicians in drug selection. Network meta-analysis can enable the comparison of multiple interventions, rank the efficacy and safety of different interventions, and thus provide the optimal treatment plan [15] .

## Methods and analysis

### Protocol and registration

We registered the protocol of this network meta-analysis on the International Prospective Register of Systematic Reviews (PROSPERO): CRD42022364242. This study will be conducted and reported according to the Preferred Reporting Items for Systematic Reviews and MetaAnalyses (PRISMA) checklist, PRISMA protocol statement, and the PRISMA-extension statement for network

meta-analysis[16] .

### Eligibility criteria

#### Study selection

The inclusion criteria were as follows:

1. Study type: Randomized controlled trials (RCTs);
2. Participants: Patients with a clear diagnostic basis for adolescent depressive disorder, elderly depressive disorder, postpartum depressive disorder, or perimenopausal depressive disorder [17] [18] **Error! Reference source not found.**;
3. Interventions:

Experimental group: Oral Chinese patent medicines combined with selective serotonin reuptake inhibitors (SSRIs). Six representative SSRIs were selected in this study, including fluoxetine, paroxetine, fluvoxamine, sertraline, citalopram, and escitalopram[19] ;

Control group: SSRIs used alone or SSRIs combined with placebo.

No restrictions were imposed on patients’ diagnosis and treatment protocols, drug doses, intervention duration, evaluation indicators, or treatment courses;

4. Outcome measure:

The Hamilton Depression Rating Scale (HAMD) was used to assess the severity of depression, with higher scores indicating more prominent depressive symptoms.

#### Data extraction

The exclusion criteria were as follows:

1. Duplicate or redundant reports, or studies confirmed to be based on the same clinical trial despite minor differences in presentation;
2. Studies with mixed disease types that make efficacy evaluation difficult;
3. Reviews, animal experiments, and theoretical research (e.g., narrative reviews, systematic reviews without meta-analysis, in vitro studies);
4. Studies without full-text access, or published in a language other than English or Chinese;
5. Studies published in non-core journals;
6. Studies where the sample size of the experimental group is less than 20 cases.

### Search strategy

The literature sources were systematically searched in the National Knowledge Infrastructure Database (CNKI), China Science Periodic Database (CSPD), Chinese Science and Technology Journal Database (CCD), China Biology Medicine (CBM), EMbase, Pubmed, Cochrane Library, and Web of Science databases, and the ClinicalTrials.gov clinical registration system was queried. The literature search was conducted until October 2023. The search strategy uses the combination of subject words and free words. The Chinese search words are depression, depression, depressive disorder, depression syndrome, depression, visceral irritability, lily disease, epilepsy, epilepsy syndrome, traditional Chinese patent medicines and simple preparations, ointment, pill, powder, granule, oral liquid, capsule, tincture, syrup, tablet, injection; The English search term is Depression; Depressive Disorder; Drugs, Chinese Herb; Chinese Traditional; Medicine, East Asian Traditional; Chinese patent drug, etc.The detailed search strategy of this study will be shown in Table 1.

**Table 1.**
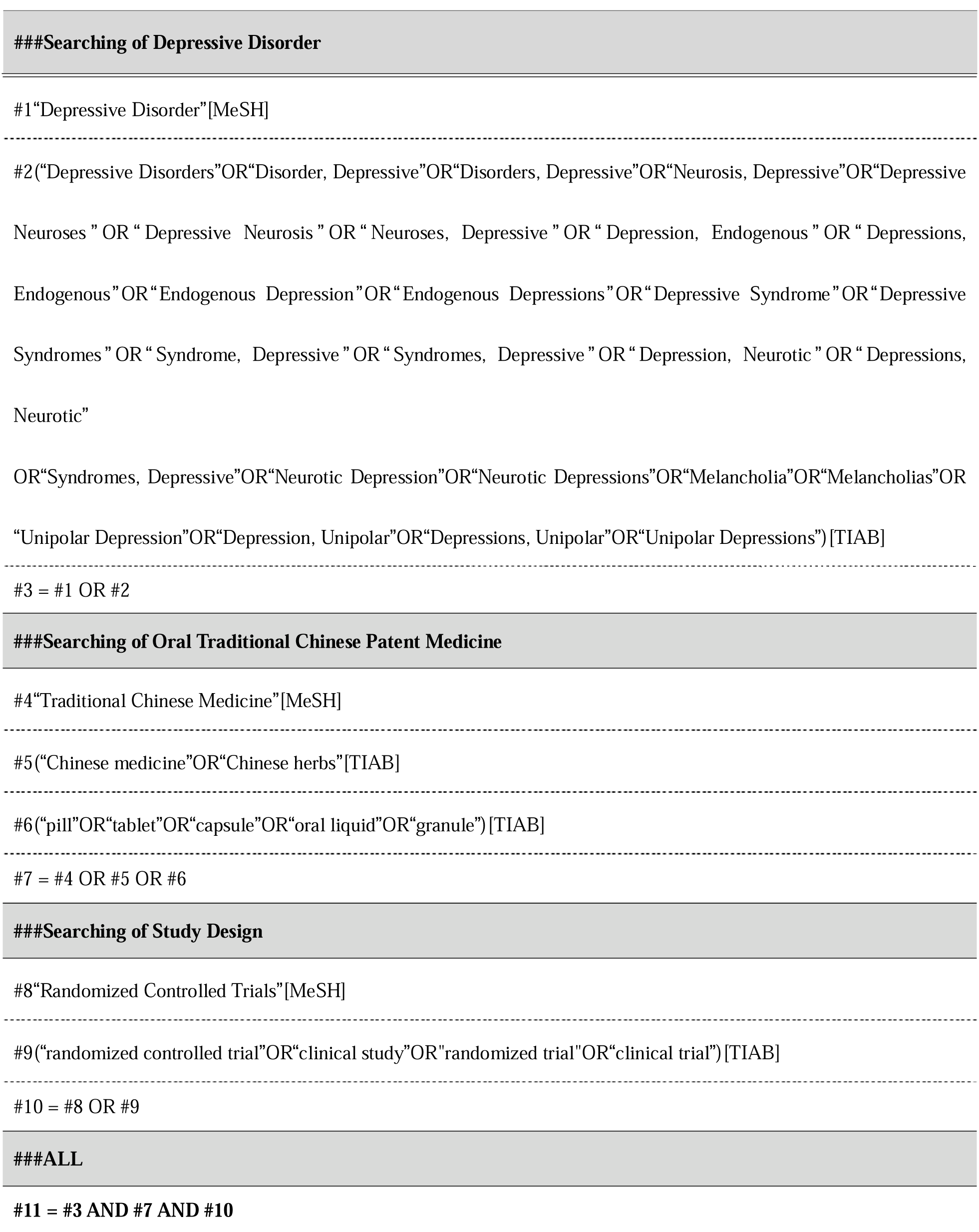
Search strategy.

### Data extraction and management

Data extraction will be conducted independently and verified by two researchers in accordance with the same criteria, with all extracted information documented in their respective data collection forms. In case of any discrepancies or ambiguities in the data, discussions with senior professionals shall be held to reach a consistent conclusion. For missing data, or vague expressions and definitions of data, communications with the corresponding authors and the publishing journals will be initiated to obtain accurate data information whenever possible. Prior to data processing and analysis, all data must undergo a re-cross-check.

The extracted data and information include the following research characteristics of participants with adolescent, elderly, postpartum and perimenopausal depression: author(s), year of publication, sample size, disease duration, treatment course, intervention measures in the treatment group, and outcome indicators [Hamilton Depression Rating Scale (HAMD) scores and adverse events].

### Grading the quality of evidence

The quality of included literature will be assessed using the Jadad Quality Scale **Error! Reference source not found.**, with the main evaluation contents covering: generation of random sequence, allocation concealment, blinding, and dropouts/withdrawals. Studies with an overall score of 1–3 points will be classified as low-quality, while those with a score of 4–7 points will be defined as high-quality.

### Statistical analysis

Network meta-analysis will be conducted using Stata 17.0 software. For continuous data, the mean difference (MD) will be used as the effect size indicator, and the 95% confidence interval (95%CI) will be adopted for interval estimation.A network plot will be generated, where the thickness of the connecting lines reflects the number of studies comparing different intervention measures, and the size of the dots corresponds to the sample size. If a closed loop is formed among the intervention measures, an inconsistency test will be performed; meanwhile, the efficacy of each intervention measure will be ranked based on the surface under the cumulative ranking curve (SUCRA) values. Finally, publication bias and small sample effects were evaluated by comparing and correcting funnel plots.

## Results

### Study selection

Tasks of screening, study selection, and data extraction will be performed independently by two reviewers. The literature will be input into the EndnoteX9 to screen the title and abstract, and the duplications and ineligible studies will be excluded. The final eligible studies will be included after reading the full text of the remaining studies. The corresponding author will be contacted if the full text is not available. Disagreements will be resolved by a third reviewer. The entire process of study selection is shown in Figure 1.

**Figure 1.**
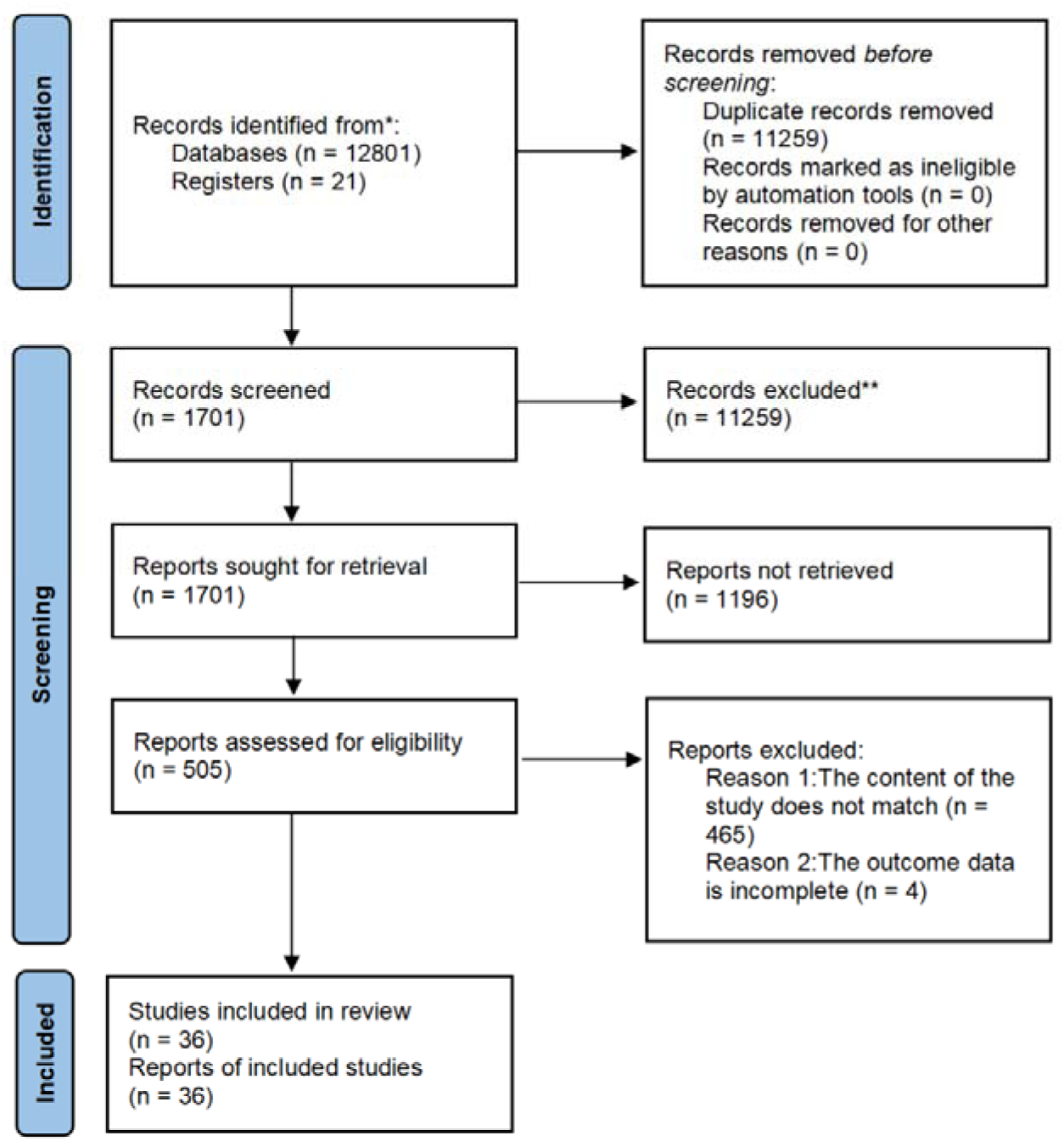
Summary of evidence search and selection.

### Characteristics of the included population

All 36 included studies were two-arm trials, involving a total of 3,110 patients with depressive disorders-1,556 patients in the experimental group and 1,554 patients in the control group. The baseline data between the two groups were comparable. The sample size of the included studies ranged from 46 to 174 cases, and four types of Chinese patent medicines were involved: Shugan Jieyu Capsules, Xiaoyao Powder, Wuling Capsules, and Morinda officinalis oligose. The baseline data of the included studies are presented in Tables 2-5.

**Table 2.**
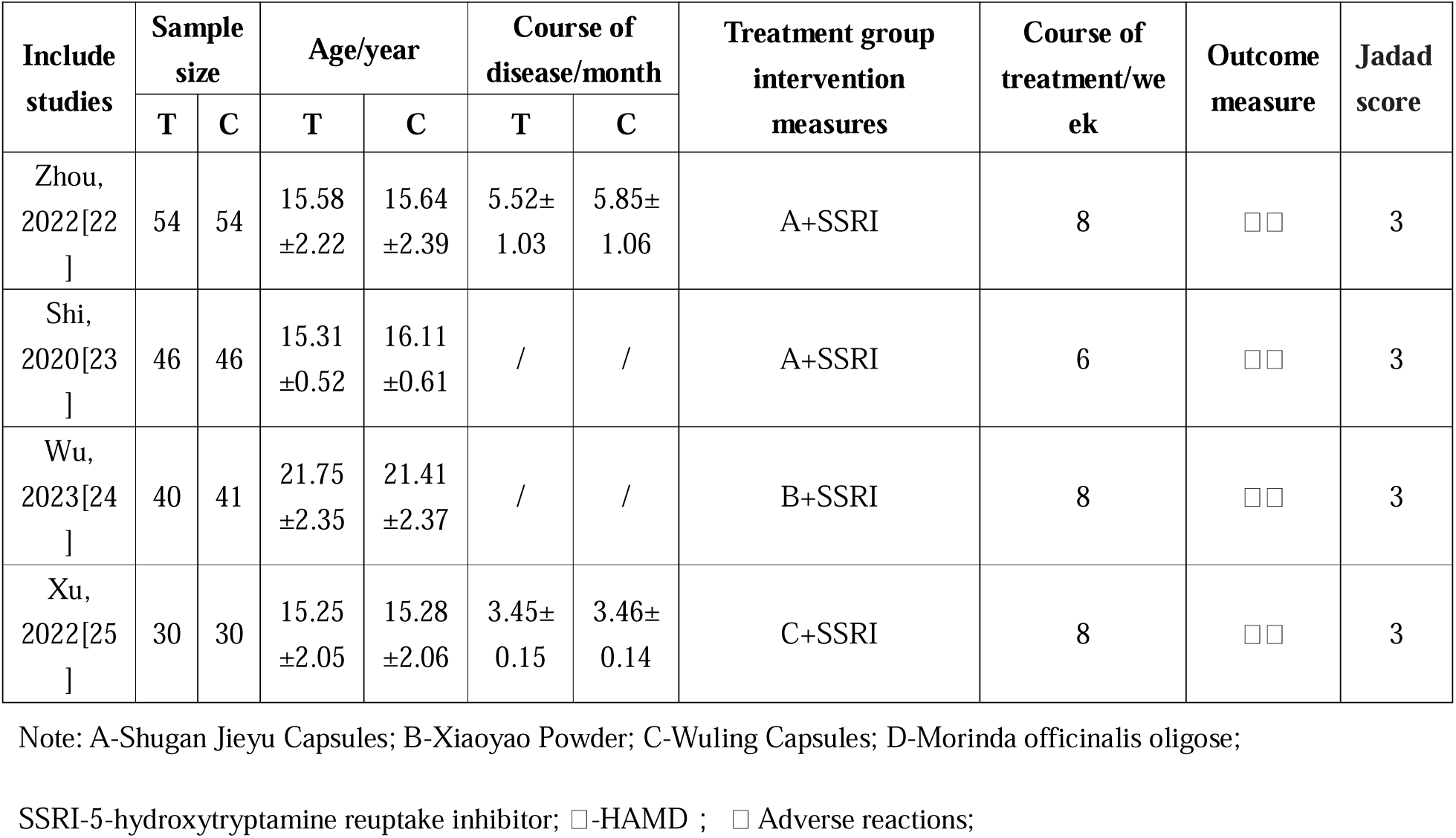
Baseline data for inclusion of adolescent depression in the study.

**Table 3.**
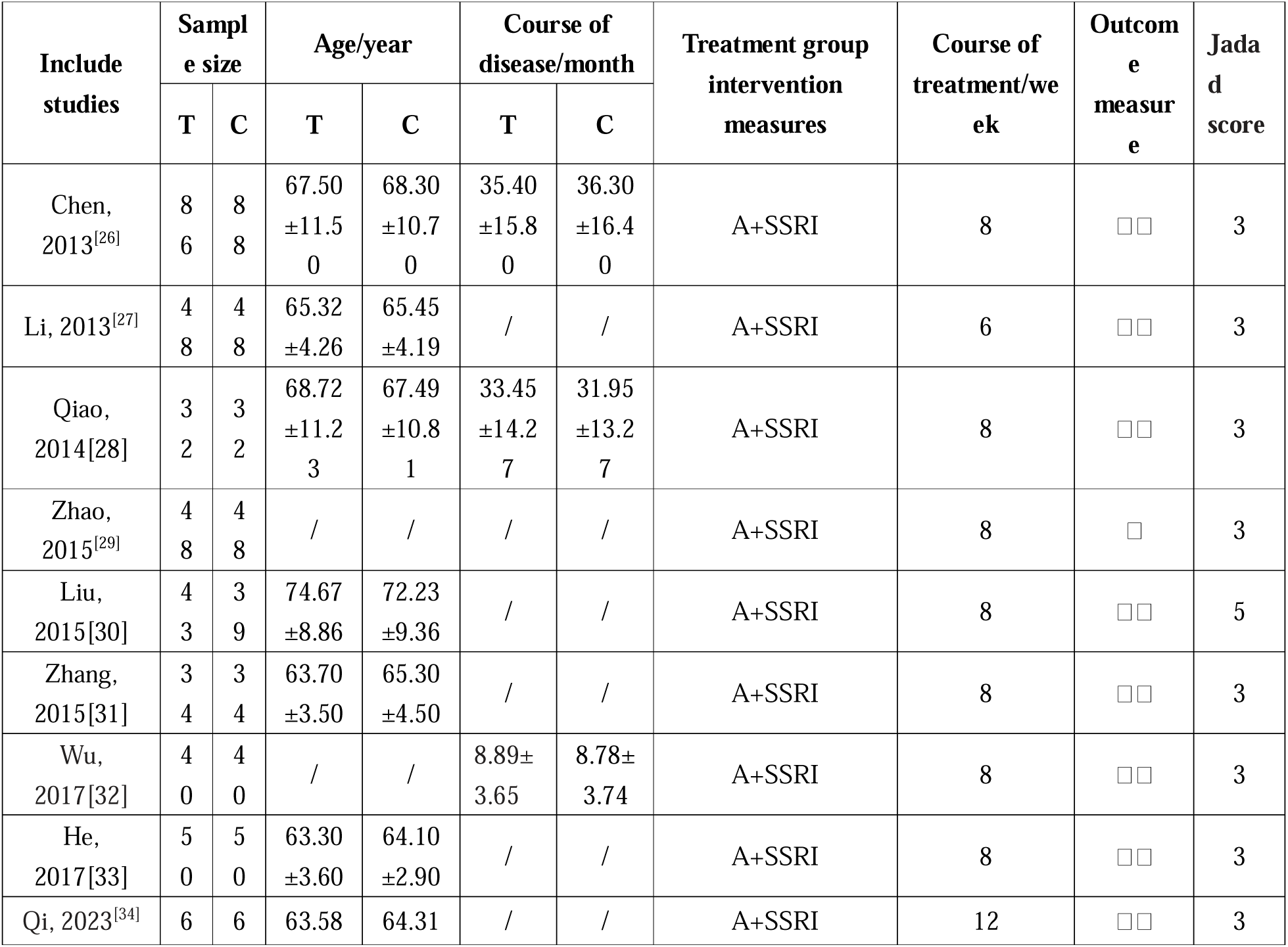
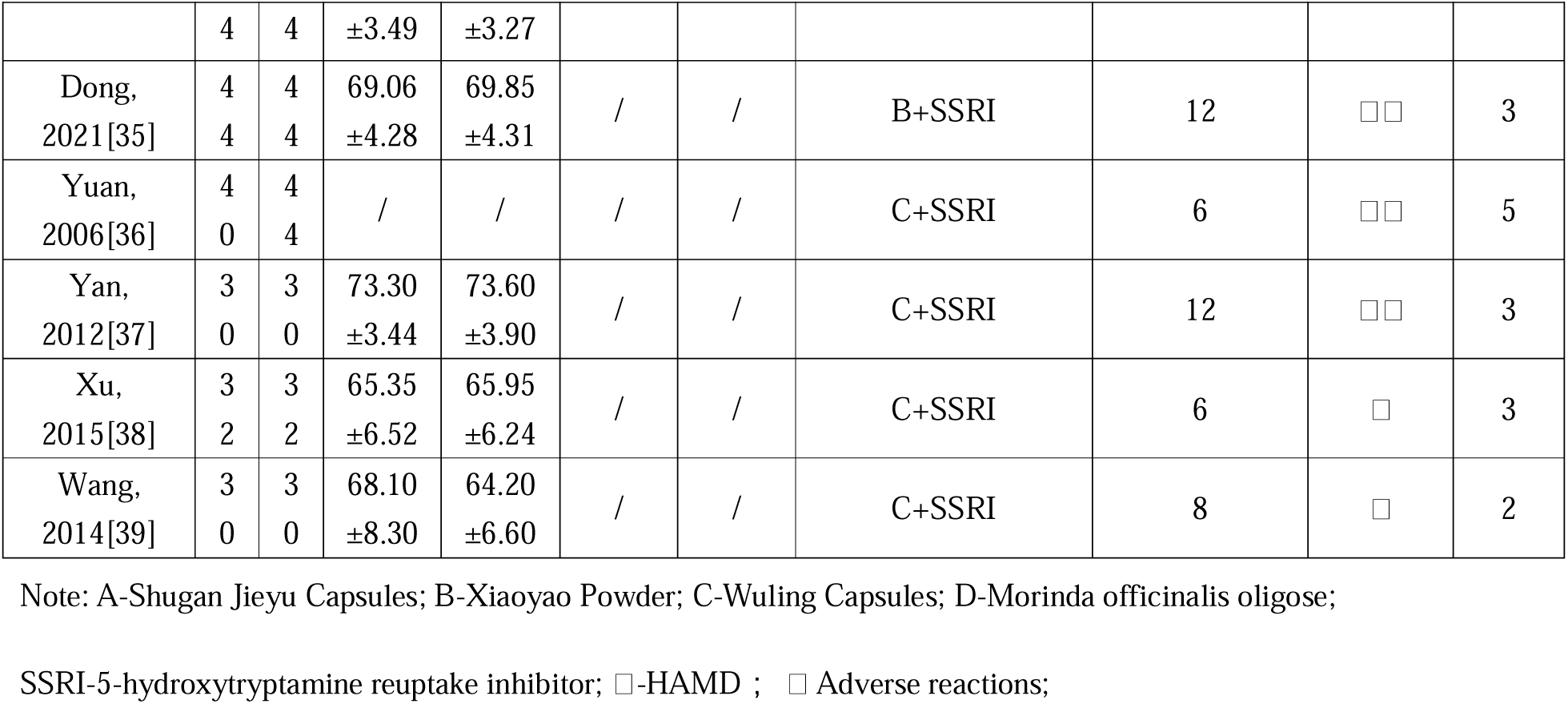
Baseline data for inclusion of elderly depression disorders in the study.

**Table 4.** Baseline data for inclusion of postpartum depression in the study.

| Include studies | Sample size |  | Age/year |  | Course of disease/month |  | Treatment group intervention measures | Course of treatment/week | Outcome measure | Jadad score |
| --- | --- | --- | --- | --- | --- | --- | --- | --- | --- | --- |
|  | T | C | T | C | T | C |  |  |  |  |
| Hao, 2015[40] | 2<br>8 | 2<br>8 | / | / | / | / | A+SSRI | 6 | □□ | 3 |
| Qiu, 2017[41] | 6<br>0 | 6<br>0 | 31.02±10.11 | 31.45±10.58 | / | / | A+SSRI | 6 | □□ | 3 |
| Chen, 2022[42] | 3<br>9 | 3<br>9 | 26.56±1.49 | 26.34±1.55 | / | / | A+SSRI | 6 | □□ | 3 |
| Liang, 2012[43] | 6<br>0 | 6<br>0 | 26.12±4.16 | 25.74±5.01 | / | / | B+SSRI | 8 | □□ | 3 |
| Li, 2016[44] | 3<br>2 | 3<br>2 | 27.58±5.14 | 26.82±4.64 | / | / | B+SSRI | 6 | □□ | 3 |
| Lin, 2020[45] | 3<br>6 | 3<br>5 | 26.98±2.30 | 27.56±2.12 | / | / | B+SSRI | 6 | □□ | 3 |
| Xie, 2023[46] | 5<br>2 | 5<br>2 | 27.56±2.12 | 27.07±3.52 | 2.54±0.62 | 2.58±0.60 | B+SSRI | 12 | □ | 2 |
| Li, 2023[47] | 4<br>8 | 4<br>8 | 29.43±2.31 | 30.87±2.15 | / | / | C+SSRI | 8 | □□ | 3 |
| Zhang, 2009[48] | 4<br>6 | 4<br>2 | 29.04±3.99 | 29.12±4.26 | / | / | C+SSRI | 6 | □□ | 3 |
| Xu, 2016[49] | 3<br>0 | 3<br>0 | / | / | / | / | C+SSRI | 6 | □□ | 2 |
| Wang, 2014[50] | 3<br>1 | 3<br>2 | 31.23±2.22 | 32.01±1.79 | / | / | C+SSRI | 4 | □ | 3 |
| Guo,<br>2022[51] | 6<br>0 | 6<br>0 | 30.20±5.5<br>0 | 30.90±5.9<br>0 | / | / | D+SSRI | 6 | □□ | 3 |

Note: A-Shugan Jieyu Capsules; B-Xiaoyao Powder; C-Wuling Capsules; D-Morinda officinalis oligose;
SSRI-5-hydroxytryptamine reuptake inhibitor; □-HAMD ; □ Adverse reactions;

**Table 5.** Baseline data for inclusion of perimenopausal depression in the study.

| Include studies | Sample size |  | Age/year |  | Course of disease/month |  | Treatment group intervention measures | Course of treatment/week | Outcome measure | Jadad score |
| --- | --- | --- | --- | --- | --- | --- | --- | --- | --- | --- |
|  | T | C | T | C | T | C |  |  |  |  |
| Hu, 2018[52] | 40 | 39 | 48.20<br>±3.70 | 48.60<br>±3.90 | / | / | B+SSRI | 8 | □□ | 2 |
| Zhang, 2011[53] | 30 | 30 | 43.50<br>±12.4<br>3 | 42.25<br>±11.3<br>8 | / | / | C+SSRI | 6 | □□ | 2 |
| Wang, 2022[54] | 50 | 50 | 49.73<br>±3.27 | 50.26<br>±3.34 | / | / | C+SSRI | 8 | □□ | 3 |

Note: A-Shugan Jieyu Capsules; B-Xiaoyao Powder; C-Wuling Capsules; D-Morinda officinalis oligose;
SSRI-5-hydroxytryptamine reuptake inhibitor; □-HAMD ; □ Adverse reactions;

### Outcome Measures

#### Evidence Network

##### 1. Adolescent Depressive Disorder

In the included analysis, a total of 4 studies (involving 3 types of Chinese patent medicines and 341 patients) reported Hamilton Depression Rating Scale (HAMD) scores. No closed loops were formed, so consistency testing was not required. The results showed that the comparison between the “Shugan Jieyu Capsules + SSRI” group and the “SSRI alone” group had the largest number of studies (2 RCTs) and the largest sample size (n=200). The network plot of the relationships between different interventions is shown in Figure 3.

**Figure 3.**
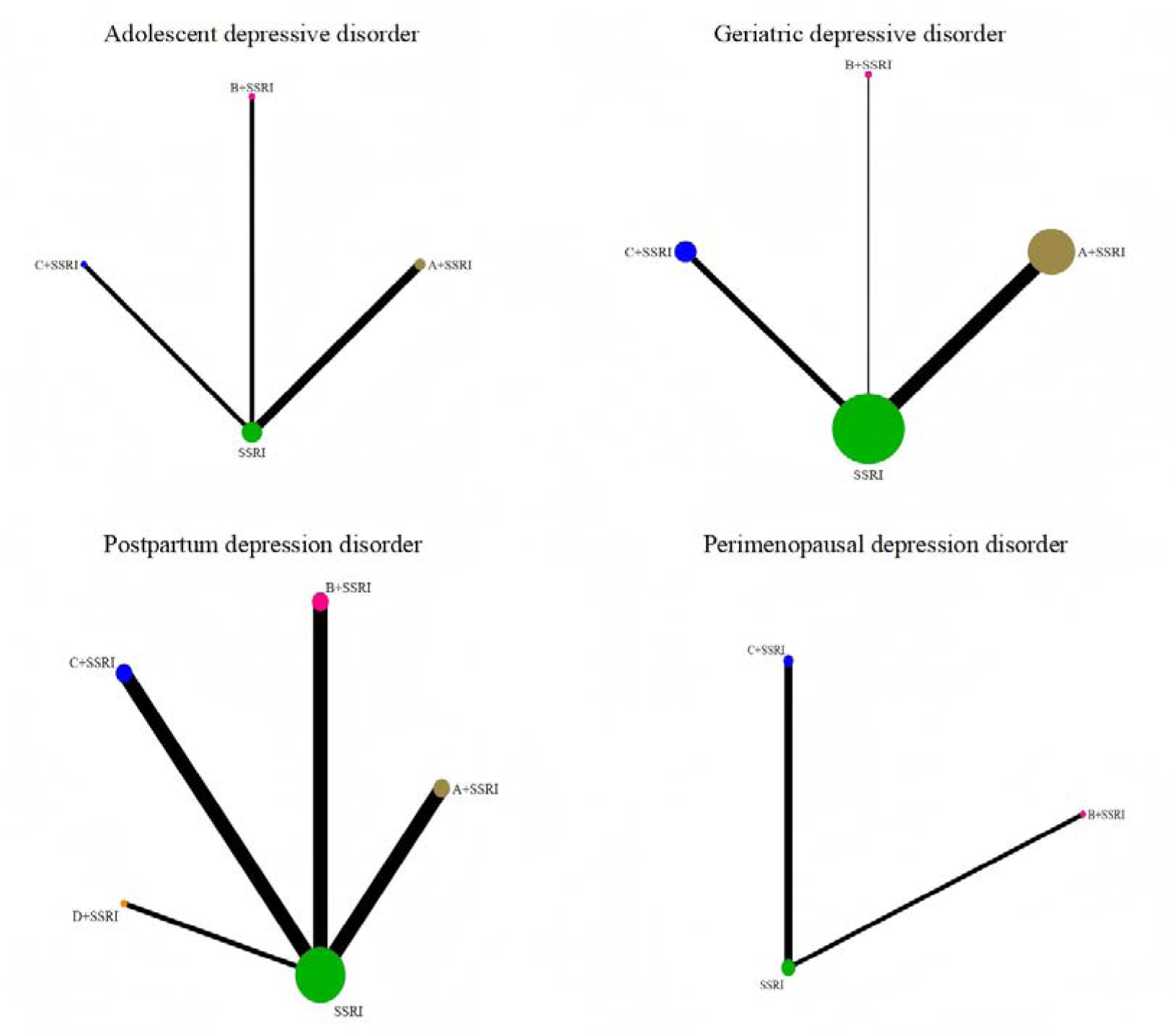
Evidence network of HAMD score. Note: A-Shugan Jieyu Capsules; B-Xiaoyao Powder; C-Wuling Capsules; D-Morinda officinalis oligose;

##### 2. Elderly Depressive Disorder

For HAMD score reporting, 14 studies were included, involving 3 types of Chinese patent medicines and 1,244 patients. No closed loops were observed, thus consistency testing was unnecessary. The findings indicated that the comparison between the “Shugan Jieyu Capsules + SSRI” group and the “SSRI alone” group had the most studies (9 RCTs) and the largest sample size (n=888). The network relationship among the interventions is presented in Figure 3.

##### 3. Postpartum Depressive Disorder

A total of 10 studies (covering 4 types of Chinese patent medicines and 873 patients) reported HAMD scores. No closed loops were formed, so consistency testing was not conducted. The results were as follows:

Comparison between “Xiaoyao Powder + SSRI” group and “SSRI alone” group: 3 RCTs, sample size n=255;

Comparison between “Wuling Capsules + SSRI” group and “SSRI alone” group: 3 RCTs, sample size n=244;

Comparison between “Shugan Jieyu Capsules + SSRI” group and “SSRI alone” group: 3 RCTs, sample size n=254.

The network plot of the relationships between the interventions is shown in Figure 3.

##### 4. Perimenopausal Depressive Disorder

For HAMD score analysis, 3 studies were included, involving 3 types of Chinese patent medicines and 239 patients. No closed loops were formed, so consistency testing was not required. The results demonstrated that the comparison between the “Wuling Capsules + SSRI” group and the “SSRI alone” group had the largest number of studies (2 RCTs) and the largest sample size (n=160). The network relationship among the interventions is displayed in Figure 3.

### Network Meta-Analysis

#### HAMD Scores

##### 1. Adolescent Depressive Disorder

Results of the network meta-analysis showed that among the 6 pairwise comparisons generated, 1 was statistically significant.The detailed is shown in Table 6. Based on the mean difference (MD) and 95% confidence interval (95%CI), it can be concluded that SSRI combined with Xiaoyao Powder was more effective than SSRI alone. The surface under the cumulative ranking curve (SUCRA) probability ranking was as follows: Xiaoyao Powder + SSRI (94.2%) > Wuling Capsules + SSRI (66.4%) > Shugan Jieyu Capsules + SSRI (39.3%) > SSRI alone (0.1%) .The detailed is shown in Table 7.

**Table 6.** Network Meta analysis of HAMD Score.

| Special populations | Intervention methods | D+SSRI | C+SSRI | B+SSRI | A+SSRI |
| --- | --- | --- | --- | --- | --- |
| adolescent | C+SSRI |  |  |  |  |
|  | B+SSRI |  | 0.93(-0.84,2.70) |  |  |
|  | A+SSRI |  | -0.75(-2.23,0.72) | -1.68(-3.31,-0.06) |  |
|  | SSRI |  | -2.00(-3.16,-0.84) | -2.93(-4.27,-1.59)* | -1.25(-2.16,-0.34) |
| elderly | C+SSRI |  |  |  |  |
|  | B+SSRI |  | 0.39(-3.05,3.82) |  |  |
|  | A+SSRI |  | 0.68(-1.36,2.73) | 0.30(-2.89,3.48) |  |
|  | SSRI |  | -2.19(-3.90,-0.49)* | -2.58(-5.56,0.40) | -2.88(-4.01,-1.75)* |
| postpartum | D+SSRI |  |  |  |  |
|  | C+SSRI | 0.48(-2.82,3.78) |  |  |  |
|  | B+SSRI | 0.07(-3.21,3.35) | -0.41(-2.89,2.08) |  |  |
|  | A+SSRI | 0.83(0.15,4.46) | 0.93(-1.55,3.41) | 1.34(-1.14,3.81) |  |
|  | SSRI | 1.41(-1.88,4.69) | -3.06(-4.83,-1.29)* | -2.65(-4.39,-0.91)* | -3.99(-5.74,-2.24)* |
| perimenopause | C+SSRI |  |  |  |  |
|  | B+SSRI |  | -1.72(-7.58,4.14) |  |  |
|  | SSRI |  | -3.12(-6.60,0.36) | -1.40(-6.12,3.32) |  |

Note: A-Shugan Jieyu Capsules; B-Xiaoyao Powder; C-Wuling Capsules; D-Morinda officinalis oligose; ( \* $p < 0.05$ )

**Table 7.**
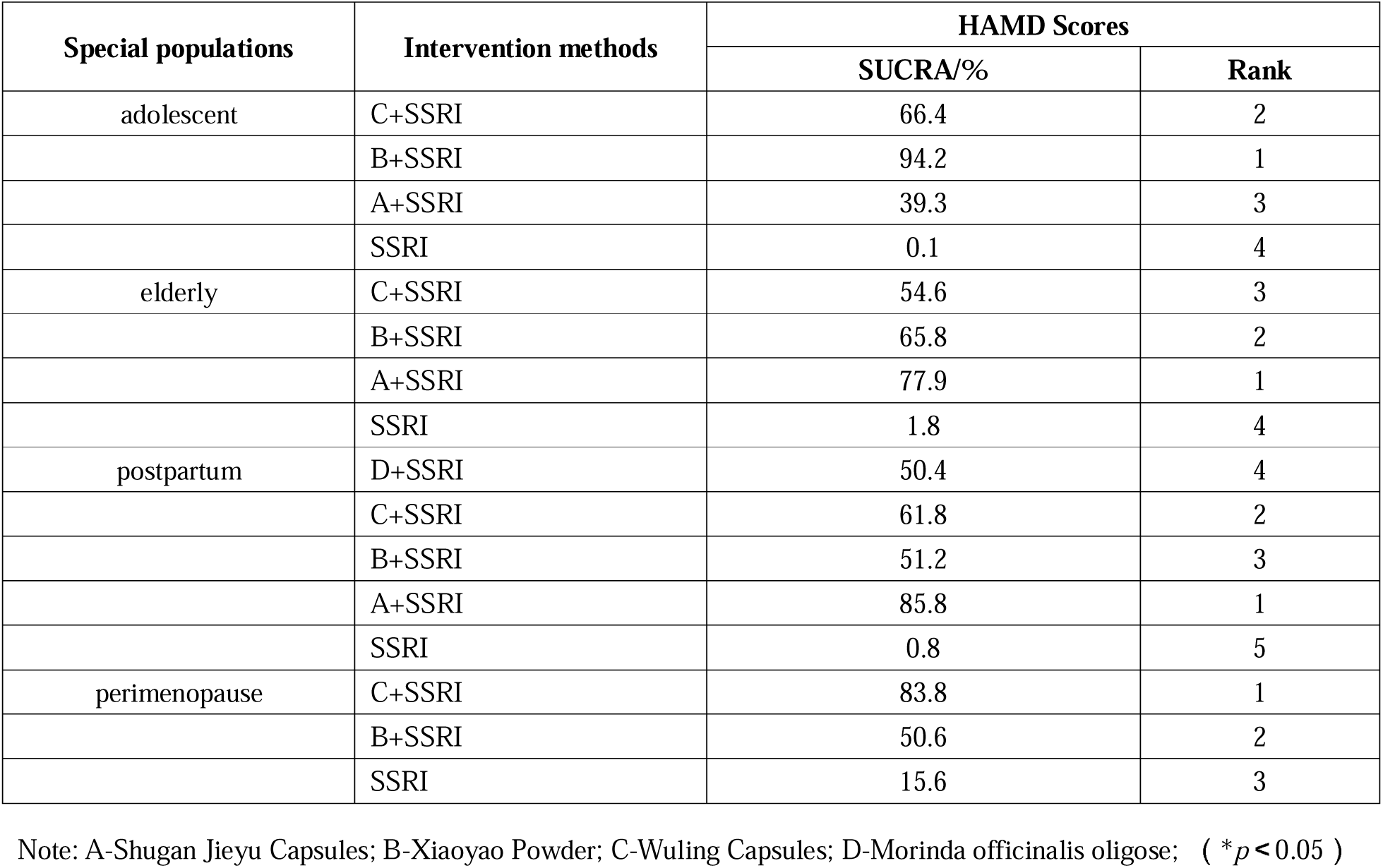
Ranking of SUCRA values based on HAMD rating.

##### 2. Elderly Depressive Disorder

Network meta-analysis results indicated that 2 out of the 6 pairwise comparisons generated were statistically significant.The detailed is shown in Table 6. According to the MD and 95%CI, SSRI combined with either Wuling Capsules or Shugan Jieyu Capsules exhibited better efficacy than SSRI alone. The SUCRA probability ranking was: Shugan Jieyu Capsules + SSRI (77.9%) > Xiaoyao Powder + SSRI (65.8%) > Wuling Capsules + SSRI (54.6%) > SSRI alone (1.8%).The detailed is shown in Table 7.

##### 3. Postpartum Depressive Disorder

Among the 10 pairwise comparisons generated by the network meta-analysis, 3 were statistically significant.The detailed is shown in Table 6. Based on the MD and 95%CI, it was found that SSRI combined with Wuling Capsules, Xiaoyao Powder, or Shugan Jieyu Capsules was more effective than SSRI alone. The SUCRA probability ranking was: Shugan Jieyu Capsules + SSRI (85.8%) > Wuling Capsules + SSRI (61.8%) > Xiaoyao Powder + SSRI (51.2%) > Morinda officinalis oligose + SSRI (50.4%) > SSRI alone (0.8%) .The detailed is shown in Table 7.

##### 4. Perimenopausal Depressive Disorder

Network meta-analysis results showed that none of the 6 pairwise comparisons generated were statistically significant.The detailed is shown in Table 6. According to the MD and 95%CI, SSRI combined with Wuling Capsules or Xiaoyao Powder had similar efficacy to SSRI alone. The SUCRA probability ranking was: Wuling Capsules + SSRI (83.8%) > Xiaoyao Powder + SSRI (50.6%) > SSRI alone (15.6%) .The detailed is shown in Table 7.

### Publication bias

The calibration comparison funnel plot displays two different intervention measures for direct comparison, represented by dots of different colors. The calibration comparison funnel plot has poor symmetry, indicating possible publication bias and small sample effects, as shown in Figure 4.

**Figure 4.**
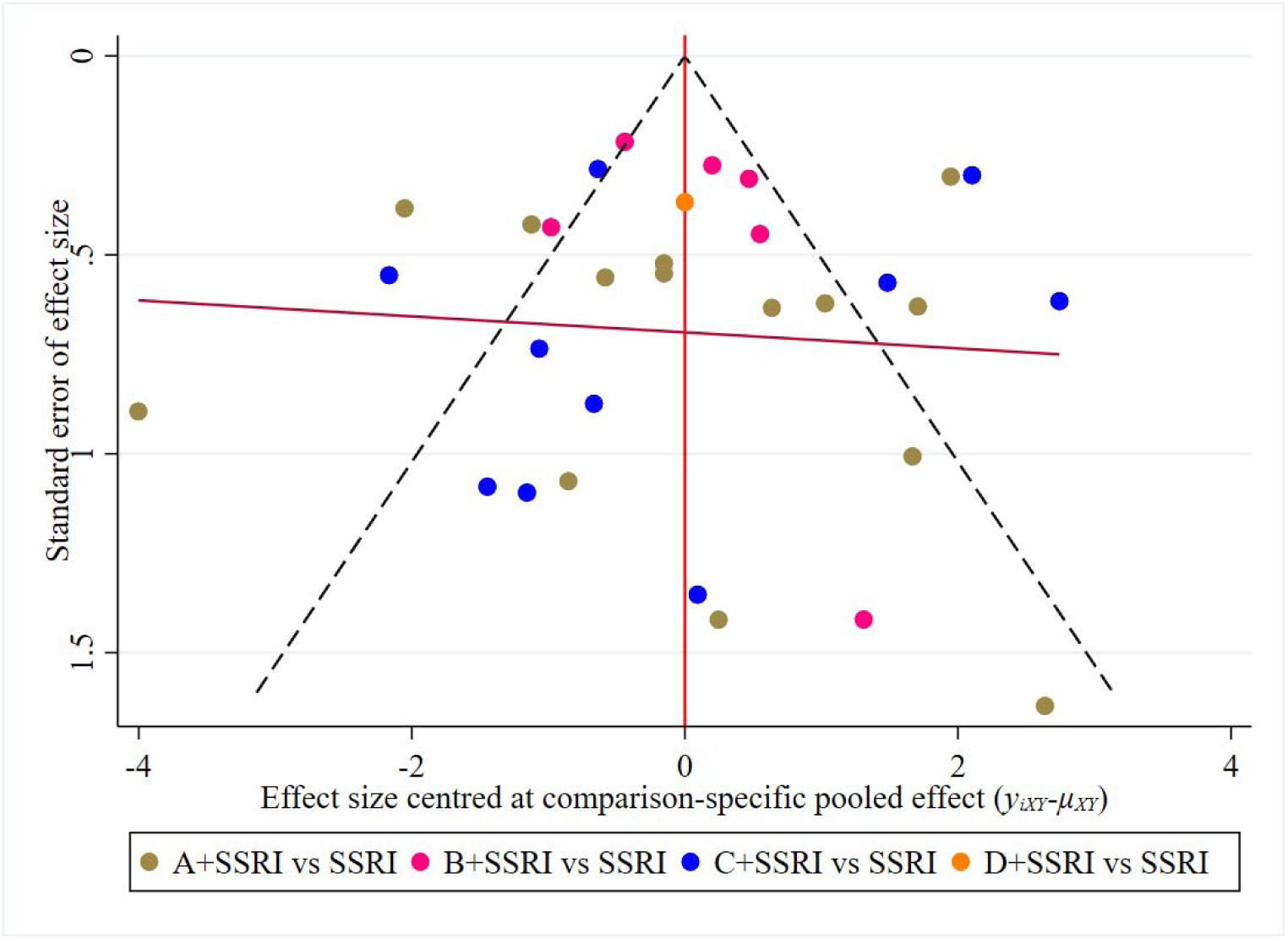
HAMD Scoring Funnel Chart. Note: A-Shugan Jieyu Capsules; B-Xiaoyao Powder; C-Wuling Capsules; D-Morinda officinalis oligose;

### Occurrence of Adverse Reactions/Events

27 studies reported adverse reactions, as shown in Table 8.

**Table 8.** Occurrence of Adverse Reactions.

| Include studies | Intervention in Traditional Chinese Medicine | Adverse reactions |  |
| --- | --- | --- | --- |
|  |  | treatment group | control group |
| Zhou, 2022[22] | Shugan Jieyu Capsules | 1 case of dizziness, 1 case of nausea, 2 cases of diarrhea, 1 case of constipation, and 2 cases of loss of appetite | 3 cases of dizziness, 2 cases of nausea, 3 cases of diarrhea, 3 cases of constipation, and 5 cases of loss of appetite |
| Shi, 2020[23] | Shugan Jieyu Capsules | 2 cases of dry mouth, 3 cases of dizziness, 1 case of diarrhea, 2 cases of anorexia, 1 case of insomnia, and 2 cases of drowsiness | 3 cases of dry mouth, 1 case of dizziness, 2 cases of diarrhea, 2 cases of anorexia, 3 cases of insomnia, and 2 cases of drowsiness |
| Wu, 2023[24] | Xiaoyao Powder | 1 case of drowsiness, 2 cases of constipation, and 1 case of nausea | 1 case of drowsiness, 1 case of constipation, and 2 cases of nausea |
| Chen, 2013[26] | Shugan Jieyu Capsules | 18 cases of adverse reaction symptoms, mainly including dry mouth, insomnia, diarrhea, etc | 16 cases of adverse reaction symptoms, mainly including dry mouth, insomnia, diarrhea, etc |
| Li, 2013[27] | Shugan Jieyu Capsules | The two groups of adverse reactions mainly include nausea, dry mouth, constipation, decreased appetite, frequent urination, dizziness, drowsiness, etc., which do not require special treatment |  |
| Qiao, 2014<sup>[28]</sup> | Shugan Jieyu Capsules | 1 case each of dizziness and diarrhea, 2 cases each of dry mouth, insomnia, abnormal liver function, and 4 cases of frequent urination | 2 cases each of dizziness, insomnia, and palpitations, 1 case each of T-wave changes on electrocardiogram, abnormal liver function, and frequent urination |
| Liu, 2015[30] | Shugan Jieyu Capsules | 2 cases of nausea and 1 case of dry mouth | 4 cases of dizziness, 3 cases of tachycardia, 3 cases of weight gain each, 2 cases of constipation, 2 cases of dry mouth, 2 cases of sexual dysfunction, and 1 case of blurred vision |
| Zhang, 2015[31] | Shugan Jieyu Capsules | 1 case of headache, 1 case of dizziness, 2 cases of frequent urination, 2 cases of insomnia, 2 cases of abnormal liver function, and 3 cases of nausea | 1 case of dizziness, 1 case of nausea, 1 case of frequent urination, 2 cases of insomnia, 2 cases of abnormal liver function, and 2 cases of T-wave changes on electrocardiogram |
| Wu, 2017[32] | Shugan Jieyu Capsules | 13 cases of adverse reactions such as insomnia, diarrhea, and dry mouth | 11 cases of adverse reactions such as insomnia, diarrhea, and dry mouth |
| He, 2017[33] | Shugan Jieyu Capsules | 6 cases of adverse reactions such as insomnia, headache, dizziness, and insomnia | 6 cases of adverse reactions such as insomnia, headache, dizziness, and insomnia |
| Qi, 2023[34] | Shugan Jieyu Capsules | Insomnia, dry mouth, drowsiness, fatigue, dizziness, headache, nausea and vomiting, etc | Insomnia, dry mouth, drowsiness, fatigue, dizziness, headache, nausea and vomiting |
| Dong, 2021[35] | Xiaoyao Powder | Gastrointestinal reactions | Gastrointestinal reactions |
| Yan, 2012[37] | Wuling Capsules | 2 cases of mild nausea | 1 case of mild dry mouth and insomnia |
| Hao, 2015[40] | Shugan Jieyu Capsules | 1 case of headache, 2 cases of nausea, 1 case of drowsiness, 1 case of dizziness, and 1 case of fatigue | Nausea in 3 cases, fatigue in 1 case, drowsiness in 2 cases, and blurred vision in 1 case |
| Qiu, 2017[41] | Shugan Jieyu Capsules | Dizziness and gastrointestinal reactions | Dizziness and gastrointestinal reactions |
| Chen, 2022[42] | Shugan Jieyu Capsules | Dizziness and gastrointestinal reactions | Dizziness and gastrointestinal reactions |
| Liang, 2012[43] | Xiaoyao Powder | 24 cases of adverse reactions | 43 cases of adverse reactions |
| Li, 2016[44] | Xiaoyao Powder | 13 cases of adverse reactions | 21 cases of adverse reactions |
| Lin, 2020[45] | Xiaoyao Powder | Sleepiness and gastrointestinal reactions | Sleepiness and gastrointestinal reactions |
| Xie, 2023[46] | Xiaoyao Powder | Rash, headache, nausea | Rash, headache, nausea |
| Li, 2023[47] | Wuling Capsules | 2 cases of drowsiness | 1 case of dizziness, 2 cases of nausea |
| Zhang, 2009[48] | Wuling Capsules | 3 cases had nausea and decreased appetite, 1 case had anxiety and sleep disorders | 8 cases of nausea, vomiting, and decreased appetite, 5 cases of anxiety and sleep disorders |
| Xu, 2016[49] | Wuling Capsules | 2 cases experienced adverse reactions, mainly including decreased appetite, nausea | 7 cases experienced adverse reactions, mainly including decreased appetite, nausea, vomiting, sleep disorders, etc |
| Wang, 2014[50] | Wuling Capsules | 2 cases of dizziness, 1 case of dry mouth, and 3 cases of nausea | 3 cases of dizziness, 1 case of dry mouth, 1 case of nausea, and 1 case of headache |
| Hu, 2018[52] | Xiaoyao Powder | Palpitations, gastrointestinal reactions | Palpitations, gastrointestinal reactions |
| Zhang, 2011[53] | Wuling Capsules | 5 cases of nausea, 1 case of headache, 2 cases of insomnia, and 1 case of drowsiness | 5 cases of nausea, 1 case of headache, 3 cases of insomnia, and 2 cases of drowsiness |
| Wang, 2022[54] | Wuling Capsules | gastrointestinal reactions | gastrointestinal reactions |

## Discussion

From the perspective of Traditional Chinese Medicine (TCM) theory, depression can be regarded as a syndrome complex caused by long-term liver qi stagnation, dysfunction of the spleen and stomach, and stagnation of qi and blood circulation in patients. Its core clinical manifestations are dominated by emotional disturbance, low mood, and irritability. The development of this syndrome complex reflects the profound impact of internal visceral dysfunction and disordered qi-blood circulation on mental and emotional states.According to the recommended medications in Clinical Application Guidelines for Chinese Patent Medicines in the Treatment of Depressive Disorders (2022) [19] , the commonly used Chinese patent medicines for depression in clinical practice currently include Shugan Jieyu Capsules, Wuling Capsules, Xiaoyao Pills, Shugan Granules, and Morinda officinalis oligose.

Shugan Jieyu Capsules exert the effects of soothing the liver to relieve stagnation and invigorating the spleen to calm the mind. Modern pharmacological studies **Error! Reference source not found.** have shown that Shugan Jieyu Capsules contain a variety of bioactive substances, such as hypericin, chlorogenic acid, and eleutheroside. These components block the reuptake of neurotransmitters including serotonin (5-HT), dopamine (DA), and norepinephrine (NE), promote the synthesis and expression of brain-derived neurotrophic factor (BDNF), and thereby optimize the structure and function of neuronal synapses. Meanwhile, this medication can regulate the dysfunction of the hypothalamic-pituitary-adrenal (HPA) axis, protect pheochromocytoma PC12 cells, enhance the body’s stress resistance, and improve immune function, ultimately exhibiting significant antidepressant effects.

Wuling Capsules possess the effects of tonifying the kidney, invigorating the brain, and nourishing the heart to calm the mind. Derived from the strain isolated from Ustilago esculenta (a rare medicinal fungus in China), Wuling Capsules contain adenosine, polysaccharides, sterols, various amino acids, vitamins, and trace elements. They exert antidepressant effects by increasing the brain’s uptake of glutamic acid, inhibiting the synthesis of the neurotransmitter γ-aminobutyric acid (GABA), and enhancing the binding activity of GABA receptors in the cerebral cortex [55] .

Xiaoyao Powder has the effects of soothing the liver, invigorating the spleen, nourishing blood, and regulating menstruation. Modern pharmacological studies have revealed the following: Bupleuri Radix (Chaihu) contains saikosaponins, among which saikosaponin D may regulate autophagy to participate in the formation of the NLRP3 inflammasome, further inhibiting the levels of pro-inflammatory cytokines and regulating the tryptophan-kynurenine pathway to exert antidepressant effects [57] . The combined use of Bupleuri Radix and Paeoniae Radix Alba (Baishao) can regulate the expression of multiple key target proteins such as SLC5A4, PTGS2, and CASP3, thereby participating in the metabolism of monoamine neurotransmitters and effectively exerting antidepressant effects through inhibiting the reuptake of 5-HT [58] . On the other hand, Poria (Fuling) is rich in polysaccharides and triterpenoids, which exhibit pharmacological effects such as liver protection, sedation, and enhancement of immune function [59] .

Morinda officinalis oligose exert the effects of relieving depression to calm the mind and tonifying the kidney to improve intelligence. Modern studies[60] have indicated that Morinda officinalis oligose exert antidepressant effects mainly through the serotonergic system. They may also influence neurotrophic factor pathways, regulate synaptic plasticity, increase the levels of BDNF and glial cell line-derived neurotrophic factor (GDNF), and decrease the level of vascular endothelial growth factor (VEGF) to achieve antidepressant effects.

In clinical practice, medical staff need to select the optimal therapeutic regimen among various oral antidepressant Chinese patent medicines. Compared with traditional meta-analysis, network meta-analysis can integrate and quantitatively compare the efficacy data of multiple interventions for the same disease, rank the efficacy based on different outcome indicators, and visualize the results. Therefore, this study realized the comparative ranking of antidepressant Chinese patent medicines for different special populations, proposed the optimal treatment regimens or measures, and provided references and evidence for the formulation of TCM guidelines.

Regarding the limitations of this study: 1) When evaluating these studies, we found that many lacked detailed information about randomization or blinding procedures. 2) There was a certain degree of publication bias and small sample size effects. 3) Due to our strict inclusion/exclusion criteria, the limited number of included studies may have affected the strength of evidence.

## Data Availability

All data produced in the present study are available upon reasonable request to the authors
All data produced in the present work are contained in the manuscript
All data produced are available online at

https://www.crd.york.ac.uk/prospero/

## Acknowledgments

We would like to acknowledge everyone who contributed to our report, including the authors of our references.

## Author Contributions

Conceptualization: Fuqing Ren, Xueyan Liu.

Data curation: Fuqing Ren, Xueyan Liu.

Formal analysis: Fuqing Ren, Xueyan Liu.

Investigation: Jinbang Wang.

Supervision: Lixian Cui,Jiangyan Wei.

Writing – original draft: Fuqing Ren, Xueyan Liu.

Writing – review & editing: Bin Li.

