## Supplemental Table 1. for "Network meta-analysis of oral Chinese patent medicine combined with SSRI for the treatment of depressive disorder in special population"

**Identification of studies via databases and registers**

Records removed *before screening*:

Duplicate records removed (n = 11259)

Records marked as ineligible by automation tools (n = 0)

Records removed for other reasons (n = 0)

Records identified from*:

Databases (n = 12801)

Registers (n = 21)

**Identification**

Records screened

(n = 1701)

Records excluded**

(n = 11259)

Reports sought for retrieval

(n = 1701)

Reports not retrieved

(n = 1196)

**Screening**

Reports assessed for eligibility

(n = 505)

Reports excluded:

Reason 1:The content of the study does not match (n = 465)

Reason 2:The outcome data is incomplete (n = 4)

Studies included in review

(n = 36)

Reports of included studies

(n = 36)

**Included**

*Consider, if feasible to do so, reporting the number of records identified from each database or register searched (rather than the total number across all databases/registers).

**If automation tools were used, indicate how many records were excluded by a human and how many were excluded by automation tools.

Source: Page MJ, et al. BMJ 2021;372:n71. doi: 10.1136/bmj.n71.

This work is licensed under CC BY 4.0. To view a copy of this license, visit <https://creativecommons.org/licenses/by/4.0/>
