## Supplemental Table 2. for "Network meta-analysis of oral Chinese patent medicine combined with SSRI for the treatment of depressive disorder in special population"

| **Section and Topic** | **Item #** | **Checklist item** | **Location where item is reported** |
| --- | --- | --- | --- |
| **TITLE** | | |  |
| Title | 1 | Network meta-analysis of oral Chinese patent medicine combined with SSRI for the treatment of depressive disorder in special population |  |
| **ABSTRACT** | | |  |
| Abstract | 2 | Objective  Investigating the therapeutic efficacy and safety of combined oral Chinese patent medicine with selective serotonin reuptake inhibitors (SSRIs) in special Homo sapiens populations with depressive disorders.  Methods  China National Knowledge Infrastructure(CNKI), Chinese Biomedical Literature Database(CBM), Chinese Academic Journal Database(CSPD), Chinese Science and Technology Journal Database(CCD), EMbase, Pubmed,Cochrane Library, and Web of Science were retrieved, and ClinicalTrials.gov was searched from the establishment of the database to October 2023. All included Rized controlled trials were assessed for quality using the Cochrane systematic evaluation manual.Network Meta-analysis was performed using the Stata 17.0 software.  Results  36 studies were included, and the total sample size included 3110 cases, including 1556 cases in the test group and 1554 cases in the control group. A total of Chinese patent medicines were included: Shugan Jieyu capsule, Xiaoyaosan , Wuling capsule, and Morinda officinalis oligose. The results of the network Meta-analysis showed that. Compared to antidepressant treatment with SSRI alone, Adolescent depressive disorder: In improving clinical efficiency, Wuling capsule combined with SSRI ranked the highest; In terms of improving the HAMD score, Xiaoyaosan ranked the highest; Geriatric depressive disorder: In improving clinical efficiency, Shugan Jieyu capsule combined with SSRI ranked the highest; In terms of improving the HAMD score, the Shugan Jieyu capsule combined with SSRI ranked the highest; Postpartum depression disorder: In improving clinical efficiency, the Shugan Jieyu capsule combined with SSRI ranked the highest; In terms of improving the HAMD score, the Shugan Jieyu capsule combined with SSRI ranked the highest; Perimenopausal depression disorder: In improving clinical efficiency, the Xiaoyaosan combined with SSRI ranked the highest; In terms of improving the HAMD score, the Wuling capsule combined with SSRI ranked the highest;  Conclusion  On the basis of SSRI, the addition of Chinese patent medicine can improve the effectiveness and safety of clinical treatment. It is suggested that more high-quality RCT studies of Chinese patent medicine for depression disorders, especially in adolescent depression disorder, postpartum depression disorder, perimenopausal depression disorder, are conducted to provide stronger evidence. |  |
| **INTRODUCTION** | | |  |
| Rationale | 3 | Depressive disorder refers to a group of mood disorders caused by various factors, characterized by prominent and persistent low mood as the main clinical feature. It is one of the most common mental disorders [1].Epidemiological surveys show that the annual prevalence rate of depressive disorder in China is 3.6%, and the lifetime prevalence rate is 6.8% [2]. It is estimated that by 2030, depressive disorder will surpass tumors and cardiovascular and cerebrovascular diseases to become the leading cause of global disease burden [3]. The occurrence of depressive disorder is also closely related to gender and age, among which children and adolescents, the elderly, and women are special populations prone to depressive disorder.The prevalence rate of depressive emotions among Chinese adolescents is as high as 15.4%, with a total detection rate of depressive emotions reaching 28.4% [4,5]. Depressive disorder has become the primary cause of disability and morbidity among adolescents aged 10-19 years. Selective Serotonin Reuptake Inhibitors (SSRIs) can be used for depressive disorder in children and adolescents [6]. Geriatric depression is often misunderstood as a physiological aging process, so it has not received systematic diagnosis and treatment for many years. The prevalence rate of geriatric depression in China is 15.9%, among which 36.4% of patients have mild cognitive impairment [7]. In terms of pharmacotherapy, SSRIs are the first-choice drugs [6]. Postpartum depression commonly occurs within 2 weeks after childbirth, and its symptoms can last for up to 1 year. The incidence of postpartum depression among women of childbearing age in China is 14.7%, while the incidence among elderly parturient women can be as high as 36.9% [8,9]. Currently, cognitive behavioral therapy is the most commonly used method in clinical practice for treating mild to moderate postpartum depression, and SSRIs are the first choice for severe postpartum depression [6]. Perimenopausal depressive disorder mostly occurs in women aged 45-55 years. Statistics show that the incidence rate of perimenopausal depressive disorder among Chinese women is 46% [10,11]. For mild cases, psychotherapy can be provided; for moderate to severe cases, combined use of SSRIs is recommended, and hormone replacement therapy can also effectively alleviate depressive symptoms [6].SSRIs have definite effects in clinical practice and are often recommended as first-line drugs of choice. However, they still have shortcomings such as obvious adverse reactions, withdrawal reactions, and drug resistance, leading to poor patient compliance [12]. |  |
| Objectives | 4 | Chinese patent medicines have advantages such as convenient administration, portability, and easy storage. Surveys show that approximately 70% of Chinese patent medicines are prescribed by Western medicine physicians [13]. Meanwhile, the combination of traditional Chinese medicines and chemical drugs may enhance efficacy and reduce side effects [14]. Nevertheless, due to the wide variety of such combinations and the lack of randomized controlled trials comparing different drugs, it brings inconvenience to clinicians in drug selection. Network meta-analysis can enable the comparison of multiple interventions, rank the efficacy and safety of different interventions, and thus provide the optimal treatment plan [15]. |  |
| **METHODS** | | |  |
| Eligibility criteria | 5 | The inclusion criteria were as follows:  1. Study type: Randomized controlled trials (RCTs);  2. Participants: Patients with a clear diagnostic basis for adolescent depressive disorder, elderly depressive disorder, postpartum depressive disorder, or perimenopausal depressive disorder [17-19];  3. Interventions:  Experimental group: Oral Chinese patent medicines combined with selective serotonin reuptake inhibitors (SSRIs). Six representative SSRIs were selected in this study, including fluoxetine, paroxetine, fluvoxamine, sertraline, citalopram, and escitalopram[20];  Control group: SSRIs used alone or SSRIs combined with placebo.  No restrictions were imposed on patients’ diagnosis and treatment protocols, drug doses, intervention duration, evaluation indicators, or treatment courses;  4.Outcome measure:  The Hamilton Depression Rating Scale (HAMD) was used to assess the severity of depression, with higher scores indicating more prominent depressive symptoms.  The exclusion criteria were as follows:  1. Duplicate or redundant reports, or studies confirmed to be based on the same clinical trial despite minor differences in presentation;  2. Studies with mixed disease types that make efficacy evaluation difficult;  3. Reviews, animal experiments, and theoretical research (e.g., narrative reviews, systematic reviews without meta-analysis, in vitro studies);  4. Studies without full-text access, or published in a language other than English or Chinese;  5. Studies published in non-core journals;  6. Studies where the sample size of the experimental group is less than 20 cases. |  |
| Information sources | 6 | The literature sources were systematically searched in the National Knowledge Infrastructure Database (CNKI), China Science Periodic Database (CSPD), Chinese Science and Technology Journal Database (CCD), China Biology Medicine (CBM), EMbase, Pubmed, Cochrane Library, and Web of Science databases, and the ClinicalTrials.gov clinical registration system was queried. The literature search was conducted until October 2023. |  |
| Search strategy | 7 | The search strategy uses the combination of subject words and free words. The Chinese search words are depression, depression, depressive disorder, depression syndrome, depression, visceral irritability, lily disease, epilepsy, epilepsy syndrome, traditional Chinese patent medicines and simple preparations, ointment, pill, powder, granule, oral liquid, capsule, tincture, syrup, tablet, injection; The English search term is Depression; Depressive Disorder；Drugs，Chinese Herb；Chinese Traditional；Medicine，East Asian Traditional；Chinese patent drug, etc |  |
| Selection process | 8 | Data extraction will be conducted independently and verified by two researchers in accordance with the same criteria, with all extracted information documented in their respective data collection forms. |  |
| Data collection process | 9 | In case of any discrepancies or ambiguities in the data, discussions with senior professionals shall be held to reach a consistent conclusion. For missing data, or vague expressions and definitions of data, communications with the corresponding authors and the publishing journals will be initiated to obtain accurate data information whenever possible. Prior to data processing and analysis, all data must undergo a re-cross-check. |  |
| Data items | 10a | The main outcome measures were the Hamilton Depression Scale (HAMD) score (to assess the severity of depression, with higher scores indicating more prominent symptoms) and the occurrence of adverse reactions (to record the types and number of adverse reactions in the trial group and control group in each study).Collect all results related to HAMD score (score changes at different time points) and adverse reactions included in the study, without selective collection. The "Outcome Measures" and "Occurrence of Adverse Reactions/Events" sections of the document detail the relevant results data for each population and each intervention. |  |
|  | 10b | The collected variables include basic study information (author, publication year), subject characteristics (sample size, age, disease duration), intervention measures (types of proprietary Chinese medicines and SSRI classes in the trial group, control group intervention methods, treatment duration), outcome indicators (HAMD scores, adverse reactions), and research quality evaluation metrics (Jadad score). The "Table 2-5" (Baseline Data Table for Including Different Special Populations) in the document comprehensively lists all these variable details.For the data such as age and disease course missing in some studies, the document did not explicitly put forward hypotheses, but only mentioned "try to contact the corresponding author and the journal to obtain accurate data" during the data collection process. If it could not be obtained, it was marked with "/" in the table without additional hypothesis supplementation. |  |
| Study risk of bias assessment | 11 | The "Evidence Quality Grading" section specifies that the Jadad Quality Scale was used to evaluate the quality of included studies. The assessment covered random sequence generation, concealment of assignments, implementation of blinding, and dropout/withdrawal rates. The evaluation was independently conducted by research team members (without specifying exact numbers but emphasizing independent assessment principles). The "Jadad score" column in Table 2-5 of the document displays quality ratings for each study (1-3 points indicate low quality, 4-7 points high quality). Most studies received a 3-point rating, with a few receiving 2 or 5 points. |  |
| Effect measures | 12 | 1. HAMD Score (Continuous Data): The mean difference (MD) was used as the effect measure, with 95% confidence intervals (95% CI) calculated to compare HAMD score differences among intervention groups. The "Table 6. Network Meta-analysis of HAMD Score" provides detailed MD and 95% CI values for comparative analyses across populations. 2. Adverse Reactions (Count Data): Qualitative and quantitative analysis were conducted by describing both the number and types of adverse reactions. No risk ratio (RR) or other effect measures were explicitly used; instead, the "Table 8. Occurrence of Adverse Reactions" directly presents adverse reaction occurrences between study groups and control groups. |  |
| Synthesis methods | 13a | The synthesis was conducted based on inclusion criteria (Item 5), specifically targeting studies meeting the following requirements: those with randomized controlled trial (RCT) designs, involving specific population groups (adolescents, elderly, postpartum women, and perimenopausal depression patients), using traditional Chinese medicine combined with selective serotonin reuptake inhibitors (SSRIs) versus SSRIs alone or SSRIs combined with placebo, and containing HAMD scores or adverse reaction data. Synthesis analyses were performed separately for each population group (adolescents, elderly, postpartum women, and perimenopausal depression patients). The "Results of Syntheses" section in the document presents categorized synthesis outcomes by population group, clearly specifying the number of studies included and the types of interventions analyzed. |  |
|  | 13b | We should contact the corresponding author to obtain the missing data as far as possible, and if it cannot be obtained, it will not be included in the relevant analysis and data conversion operation will not be performed. |  |
|  | 13c | Results are presented through multiple formats: 1. Tables: "Table 6" displays the mesh meta-analysis effect sizes for HAMD scores, "Table 7" shows SUCRA rankings based on HAMD scores, and "Table 8" presents adverse reaction occurrences; 2. Figures: "Figure 3. Evidence Network of HAMD Score" illustrates comparative relationships between interventions using a network diagram (node size represents sample size, line thickness indicates study quantity), while "Figure 4. HAMD Scoring Funnel Chart" evaluates publication bias through a funnel plot, visually demonstrating results. |  |
|  | 13d | 1. Selection rationale for synthesis methods: As the study involved multiple interventions (four traditional Chinese patent medicines combined with SSRI + SSRI monotherapy), conventional Meta analysis could not effectively compare multiple interventions. Therefore, a network Meta analysis was adopted to simultaneously evaluate and rank therapeutic efficacy of different interventions (as explained in reference [15] regarding the advantages of network Meta analysis). 2. Meta analysis details: The network Meta analysis was conducted using Stata 17.0 software; no explicit mention was made of fixed or random effects models; no statistical heterogeneity test was performed (since there was no closed-loop comparison of interventions across populations in included studies, the document states "no closed-loop, therefore no consistency test required," which indirectly indicates the absence of heterogeneity testing). |  |
|  | 13e | 1. Subgroup analysis (Subgroup Analysis) is the most basic and commonly used method, which is suitable for exploring the heterogeneity of categorical variables (such as study type, gender, intervention dose).2. Grouped Forest Plot A visual tool for subgroup analysis. It helps to determine the source of heterogeneity by visually comparing the distribution of effect sizes within subgroups. |  |
|  | 13f | Excluding low quality research, changing the effect model and so on. |  |
| Reporting bias assessment | 14 | The "Publication Bias" section outlines methods for assessing publication bias and small sample effects through corrected funnel plot analysis. Figure 4 presents a funnel plot of HAMD scores, where colored dots represent different intervention comparisons (e.g., A+SSRI vs SSRI, B+SSRI vs SSRI). The text notes that "poor funnel plot symmetry suggests potential publication bias and small sample effects," emphasizing the importance of visual assessment in research design. |  |
| Certainty assessment | 15 | Document uses the Jadad scale to assess the quality of inclusion (item 11) |  |
| **RESULTS** | | |  |
| Study selection | 16a | The "Study Selection" section and "Figure 1" provide detailed information as follows: 1. Search Results: The number of retrieved articles from various databases included PubMed (4 articles), CNKI (3,007 articles), EMBASE (153 articles), CSPD (2,713 articles), Cochrane Library (42 articles), CCD (2,332 articles), Web of Science (12 articles), CBM (4,676 articles), and ClinicalTrials.gov (21 articles). 2. Screening Process: After excluding 11,259 duplicate articles, 1,196 irrelevant articles were filtered through title and abstract screening. Further removal of 465 articles (due to mismatched study content) and 4 articles (due to incomplete outcome data) was conducted during full-text review. 3. Final Results: A total of 36 studies were included. Figure 1 comprehensively illustrates the entire process flowchart. |  |
|  | 16b | Refers to excluding "research content mismatch" in Figure 1. |  |
| Study characteristics | 17 | The baseline data of the included studies are presented in Tables 2-5. |  |
| Risk of bias in studies | 18 | This study employed the Jadad Quality Scale to conduct a bias risk assessment for all 36 randomized controlled trials (RCTs) included in the analysis. The evaluation dimensions encompassed random sequence generation, allocation concealment, blinding implementation, and dropout/withdrawal rates, with final quantification of research quality using a "Jadad score" system (ranging from 1-3 points for low-quality studies to 4-7 points for high-quality ones). The bias risk assessment results for each study were categorized by special population groups, with specific data sourced from corresponding baseline data tables in the documentation (Tables 2-5). |  |
| Results of individual studies | 19 | The study categorized participants into four special groups: adolescents, elderly individuals, postpartum women, and perimenopausal women. Statistical data and effect estimates for each included outcome measure (HAMD score, adverse reactions) were presented using structured tables. All data were sourced from Tables 2 to 8 in the document and corresponding study descriptions, with no references to external materials. |  |
| Results of syntheses | 20a | According to the synthesis analysis of four special groups (adolescent depression, elderly depression, postpartum depression and perimenopausal depression), the characteristics and risk of bias of the included studies were summarized respectively. |  |
|  | 20b | The results of network Meta analysis (the core statistical synthesis method of this study) were presented for four special groups: adolescents, elderly, postpartum, and perimenopausal depression. The summary effect estimates, 95% confidence intervals (95% CI), and direction of effects for each intervention comparison were included. Due to the lack of closed-loop data, no statistical heterogeneity indicators were reported. |  |
|  | 20c | This study conducted a network Meta-analysis of depression treatment in specific populations using oral Chinese patent medicines combined with SSRIs. Unlike conventional approaches, it omitted traditional heterogeneity testing (e.g., I²-value calculation) as closed-loop intervention comparisons were unavailable across study groups. Through descriptive analysis of baseline characteristics, treatment regimens, and outcome evaluations in included studies, the research indirectly identified potential sources of heterogeneity. |  |
|  | 20d | This study did not conduct any sensitivity analysis based on the results of a network Meta-analysis of depression treatment in special populations with oral proprietary Chinese medicine combined with SSRI. |  |
| Reporting biases | 21 | Report bias risk due to missing outcomes was assessed separately for four special populations (adolescents, elderly, postpartum, and perimenopausal depression) based on synthesis analysis (all around "HAMD score" and "adverse reaction" outcomes) |  |
| Certainty of evidence | 22 | The core evaluation endpoints of this study were Hamilton Depression Rating Scale (HAMD) scores (primary efficacy endpoint) and adverse event incidence (safety endpoint). Evidence certainty for both endpoints was assessed across four specialized population groups: adolescents, elderly individuals, postpartum women, and perimenopausal depression patients. A four-tier grading framework (high, medium, low, very low) was applied (referencing GRADE principles while incorporating dimensions such as research quality, consistency, precision, and publication bias). |  |
| **DISCUSSION** | | |  |
| Discussion | 23a | I. Efficacy Results: Consistent with Existing Evidence in Integrated Chinese-Western Medicine for Depression Treatment This study demonstrates that adding specific traditional Chinese patent medicines (TCM) to SSRI-based treatment can improve depressive symptoms in various patient groups (as indicated by reduced HAMD scores), which aligns closely with current evidence in integrated TCM-Western medicine depression management. The findings are specifically reflected in the following aspects: 1. Consistency in Therapeutic Efficacy of Shugan Jieyu Capsules In this study, the combination of Shugan Jieyu Capsules and SSRI showed optimal efficacy in elderly depression (HAMD score improvement MD=-2.88,95% CI: -4.01, -1.75) and postpartum depression (MD=-3.99,95% CI: -5.74, -2.24), ranking first in SUCRA efficacy rankings. This conclusion is consistent with the findings of literature [55] (Network Pharmacology Research) —— The components in Shugan Jieyu Capsules, including golden wattle glycosides and chlorogenic acid, can block the reuptake of neurotransmitters such as 5-HT and DA while promoting BDNF expression, thereby enhancing SSRI's antidepressant effects. Additionally, the "Clinical Application Guidelines for TCM in Depression Treatment (2022)" [20] explicitly recommends Shugan Jieyu Capsules as a core medication in integrated TCM-Western medicine depression treatment, further supporting the reliability of this study's findings. The Efficacy Complementarity of Xiaoyao San and Wu Ling Capsules This study demonstrates that Xiaoyao San combined with selective serotonin reuptake inhibitors (SSRIs) shows potential advantages in adolescent depression (MD=-1.68,95% CI: -3.31, -0.06) and perimenopausal depression (SUCRA=50.6%), while Wu Ling Capsules outperform SSRIs in perimenopausal depression (SUCRA=83.8%). These findings align with existing pharmacological research conclusions: Literature [57] indicates that Chaihu Saponin D in Xiaoyao San enhances SSRI efficacy by inhibiting NLRP3 inflammatory bodies and regulating the tryptophan-cornualine pathway, particularly suitable for adolescents (where inflammation is closely associated with depression) and perimenopausal populations (where hormonal fluctuations trigger inflammatory activation). Literature [56] reveals that adenosine and polysaccharides in Wu Ling Capsules enhance cerebral cortex GABA receptor activity, synergizing with SSRIs to improve sleep-related depressive symptoms. Given that perimenopausal depression patients often experience sleep disorders, the efficacy of Wu Ling Capsules becomes more pronounced, corroborating this study's findings. Comparison with SSRI Monotherapy Current evidence suggests that SSRi monotherapy for depression in specific populations has an effective rate of approximately 50%-60%, with delayed onset (2-4 weeks) and adverse reaction rates ranging from 20%-30% [6,12]. In this study, the improvement in HAMD scores with the combination of Chinese patent medicines and selective serotonin reuptake inhibitors (SSRIs) (mean difference in treatment effects, MD: -1.68 to-3.99) was significantly higher than that of SSRIs alone (SUCRA <2%). Among 27 studies reporting adverse reactions, most indicated fewer cases of adverse reactions in the combined treatment group compared to the SSRIs-only group (e.g., Liang et al., 2012 [43]:24 cases vs. 43 cases in the SSRIs-only group). This demonstrates that the combined regimen outperforms SSRIs alone in both "enhanced efficacy" and "reduced adverse reactions", aligning with the conclusion from literature [14] that integrated Chinese and Western medicine can synergistically enhance therapeutic effects while reducing toxicity. II. Safety Results: Verification of Traditional Chinese Medicine's Adverse Reaction Mitigation Effects on SSRIs Although this study did not conduct quantitative Meta analysis of adverse reactions, descriptive data indicate that the TCM + SSRI group primarily experienced mild gastrointestinal reactions (nausea, diarrhea) and neurological responses (dizziness, drowsiness), with no severe cases reported. Most studies demonstrate lower incidence rates compared to SSRI-only groups, a finding consistent with existing safety evidence. Specifically regarding Shugan Jieyu Capsules' efficacy in reducing SSRI-related gastrointestinal reactions: Literature [55] highlights that the cordycepin in Shugan Jieyu Capsules regulates hypothalamic-pituitary-adrenal (HPA) axis function, thereby mitigating SSRI-induced gastrointestinal mucosal irritation. Our study by Zhou et al. (2022) [22] further confirms this pharmacological mechanism, showing significantly fewer cases of diarrhea (2 vs. 3) and constipation (1 vs. 3) in the TCM + SSRI group compared to SSRI-only treatment (3 vs. 3). Wuling Capsules Improve SSRI-Related Sleep Disorders. Simple SSRI treatment often leads to insomnia or drowsiness (incidence rate approximately 15%-20%[12]), while in this study, Yan et al. (2012)[37] reported only 2 cases of mild nausea in the Wuling Capsules + SSRI group, with no insomnia reported, compared to 1 case of insomnia in the control group. Literature[56] also confirmed that Wuling Capsules can improve SSRI-related sleep disorders by increasing intracerebral glutamate uptake and regulating GABA receptor activity, consistent with the results of this study. Consistency with existing safety evidence of traditional Chinese patent medicines. The "China Depression Disorder Prevention and Treatment Guidelines (Second Edition)" [6] mentioned that the incidence of adverse reactions in traditional Chinese patent medicine treatment for depression is approximately 5%-10%, significantly lower than SSRI. In this study, the number of adverse reactions in the TCM + SSRI group was generally below this level (e.g., Wu et al., 2023[24] showed only 4 cases of adverse reactions in the Xiaoyao San + SSRI group, with an incidence rate of 10%), and no severe reactions occurred, further supporting the existing evidence that "traditional Chinese patent medicines can enhance SSRI safety". III. Adaptability to Special Populations: Addressing Evidence Gaps in Integrative Medicine for Specific Groups Current integrated Chinese-Western studies on depression predominantly focus on adults, with limited evidence available for adolescents, postpartum women, and perimenopausal individuals. This research provides valuable supplementary insights: Adolescent Depression: Potential Suitability of Xiaoyao San Adolescent depression is often associated with "liver qi stagnation and emotional distress" (Traditional Chinese Medicine theory [11]), which aligns with Xiaoyao San's therapeutic effects of "regulating liver function and strengthening spleen." Our study demonstrates that Xiaoyao San combined with selective serotonin reuptake inhibitors (SSRIs) shows the highest improvement in HAMD scores among adolescent depression cases (SUCRA=94.2%), a finding previously only reported in literature [24]. Through synthesis analysis of four studies, this research provides stronger evidence for its applicability. Postpartum Depression: Validation of Shugan Jieyu Capsules' Efficacy Postpartum depression correlates with "blood and qi deficiency and liver qi stagnation," where Shugan Jieyu Capsules' functions of "regulating liver qi and alleviating depression, enhancing qi and strengthening spleen" match physiological characteristics. Our study reveals its combination with SSRIs achieves the largest effect size (MD=-3.99,95% CI: -5.74, -2.24), surpassing single-center results from literature [40] (Hao, 2015). By synthesizing data from twelve studies, this research reinforces the evidence-based rationale for this treatment protocol. IV. Perimenopausal Depression: Evidence Supplement for Wu Ling Capsules Perimenopausal depression is associated with "kidney yin deficiency and mental restlessness," and Wu Ling Capsules 'efficacy of "kidney-tonifying, brain-nourishing, heart-maintaining, and spirit-calming" aligns with this pathogenesis. This study showed that when combined with SSRI, it achieved SUCRA=83.8% improvement in HAMD scores for perimenopausal depression. Although statistically insignificant, existing studies (e.g., literature [54]) only reported single cases. Our synthesis analysis of three studies provides new reference directions for medication selection in this population. IV. Differences from Existing Evidence and Possible Reasons In this study, the combination of Morinda officinalis oligosaccharides + SSRI showed no significant efficacy in postpartum depression (MD=1.41,95% CI: -1.88,4.69), differing from literature [60] (which concluded Morinda oligosaccharides can combat depression through serotonergic systems). Possible reasons include: 1. Sample size limitation: Only one study (Guo, 2022 [51]) included a Morinda oligosaccharides + SSRI regimen with 60 cases, which may have insufficient power to detect effects; 2. Dosage discrepancy: The Morinda oligosaccharides in literature [60] was administered at 30mg daily, while this study's dosage was unspecified, potentially affecting efficacy due to inadequate dose; 3. Population differences: Literature [60] targeted general adults, whereas this study focused on postpartum populations where hormonal changes may influence drug metabolism and treatment outcomes. |  |
|  | 23b | Regarding the limitations of this study: 1) When evaluating these studies, we found that many lacked detailed information about randomization or blinding procedures. 2) There was a certain degree of publication bias and small sample size effects. 3) Due to our strict inclusion/exclusion criteria, the limited number of included studies may have affected the strength of evidence. |  |
|  | 23c | N/A |  |
|  | 23d | N/A |  |
| **OTHER INFORMATION** | | |  |
| Registration and protocol | 24a | We registered the protocol of this network meta-analysis on the International Prospective Register of Systematic Reviews (PROSPERO): CRD42022364242. |  |
|  | 24b | Registration Details: The study protocol has been registered on the International Prospective Systematic Review Registry (PROSPERO) under registration number CRD42022364242, accessible at https://www.crd.york.ac.uk/prospero/. To access the full protocol, researchers may search for "CRD42022364242" in PROSPERO's database to retrieve key components including research objectives, inclusion/exclusion criteria, search strategies, and statistical analysis plans. |  |
|  | 24c | N/A |  |
| Support | 25 | Funding:This project is jointly supported by The Beijing Hospital Management Center “peak” talent training plan team (DFL20241001). The funders had no role in study. |  |
| Competing interests | 26 | The authors have declared that no competing interests exist. |  |
| Availability of data, code and other materials | 27 | Publicly available material: The study protocol has been registered on the PROSPERO platform (number CRD42022364242) and is available through https://www.crd.york.ac.uk/prospero/queries. |  |

*From:*  Page MJ, McKenzie JE, Bossuyt PM, Boutron I, Hoffmann TC, Mulrow CD, et al. The PRISMA 2020 statement: an updated guideline for reporting systematic reviews. BMJ 2021;372:n71. doi: 10.1136/bmj.n71. This work is licensed under CC BY 4.0. To view a copy of this license, visit <https://creativecommons.org/licenses/by/4.0/>
